# Impact of FIFA World Cup 2022 on Children’s Sleep Patterns: An International Survey

**DOI:** 10.1101/2025.09.07.25335257

**Authors:** Mohamad-Hani Temsah, Fadi Aljamaan, Ibraheem Altamimi, Reem Alageel, Hadeel Alsulami, Shereen A. Dasuqi, Mohammed A. Albabtain, Amr Jamal, Shuliweeh Alenazi, Mohammed Alarabi, Kaled Saad, Sarah Alsubaie, Rabih Halwani, Fahad A. Bashiri, Khalid Alhasan, Shaikh Mohammed Iqbal, Muslim Alsaadi, Ahmed S. BaHammam

**Author notes:** Correspondance: Mohamad-Hani Temsah, MD, FRCPCH, Pediatric Department, College of Medicine, King Saud University, Riyadh, Saudi Arabia. Equally contributed first authors (Mohamad-Hani Temsah, Fadi Aljamaan).

## Abstract

**Background:** Poor sleep quality in children can lead to physical and psychosocial problems. The FIFA World Cup has been shown to impact adult behaviors, but its effect on children’s sleep patterns is less understood. The study aimed to evaluate the impact of the FIFA World Cup 2022 (FWC-2022) on children’s sleep patterns.

**Methods:** A cross-sectional survey was conducted between 27 November and 25 December 2022, targeting parents in Saudi Arabia (Arabia standard time) and countries with a +6-hour time difference. Participants completed the validated Children’s Sleep Habits Questionnaire (CSHQ), alongside demographics, time spent watching matches, and parental perceptions on sleep.

**Results:** A total of 848 parents participated, with 60.6% being mothers. The study found that children averaged 9.10 hours of sleep; 64.2% of parents observed no change, while 10.4% reported substantial changes. Parents aged ≥45 and those noticing shifts in sleep habits reported higher problematic sleep scores. Larger families reported fewer sleep issues, with a negative correlation between family size and sleep problems. Children’s CSHQ scores indicate mild to moderate sleep difficulties across domains. No significant differences were observed between Saudi Arabia and countries with +6-hour time difference. However, one-third of children experienced delays in sleep onset exceeding one hour on weekdays during the World Cup.

**Conclusion:** Sociodemographic factors, family dynamics, and major events like the FWC-2022 influence parental perceptions of child sleep issues. Older parents and smaller families reported more challenges, while higher socioeconomic status was linked to fewer bedtime difficulties. Our findings may be particularly relevant for FIFA 2026, where transcontinental hosting across North America will expose children globally to matches at even more variable times. Subtle impacts of prolonged event schedules highlight the need for interventions supporting healthy routines during such events, potentially through engaging, sleep-friendly technologies.

## 1. Introduction

Sleep plays a crucial role in children’s development, as nighttime brain processes are integral to mature cognitive and emotional development, and in turn enhances daytime performance[1]. Therefore, it is essential for healthcare providers to consistently advocate for healthy sleep habits and sufficient sleep duration, given that sleep is vital for overall well-being, including brain health especially for children’s growing brains[2]. Poor sleep quality in children has been noted to lead to physical and psychosocial problems, including obesity, anxiety and depression in adolescence[3, 4]. Additionally, recent research shows that young adolescents who commonly suffer from inadequate sleep tend to exhibit lower academic performance and diminished social-emotional skills[5].

Contemporary evidence supports the general recommendation to achieve an adequate number of sleep hours on a regular basis to promote optimal health among children. Individual variance in sleep needs is influenced by behavioral, genetic, medical, societal and environmental factors. It is recommended that children aged three to five years should sleep 10 to 13 hours daily, while children aged 6 to 12 years should sleep 9 to 12 hours per day to ensure optimal health outcomes[6].

A study explored sleep habits on school performance in elementary school children, found that watching television and using electronic devices during sleep time adversely affected school attendance and performance[7]. Additionally, a systematic review of the association between screen time (e.g., television, computers, video games, and mobile devices) and sleep outcomes among school-aged children and adolescents found that screen time adversely affects sleep outcomes (primarily shortened duration and delayed timing)[8], children and adolescents who spent more time on screens slept less hours and were more likely to get insufficient sleep duration. The associations with sleep duration were primarily due to portable electronic devices, with the importance of non-portable devices diminishing in children over age 10. These findings remained consistent even when accounting for demographic variables, diagnoses of anxiety or depression, physical activity, and BMI[9].

More recent data suggests that electronic screen exposure remains very high, one randomized clinical trial reporting that children averaged nearly 36 hours per week of recreational screen use at baseline, more than 5 hours per day. This underscores how central electronic media usage in daily life and their impact on sleep continues to be a concern[10]. Furthermore, during the COVID-19 lockdown, the lifestyle of children and adolescents was profoundly disrupted, with limited peer interaction, reduced physical activity, and less exposure to natural daylight. Confinement at home also led to a sharp increase in the use of electronic devices, both for leisure and for school activities as distance learning replaced in-person lessons. This greater reliance on screens introduced additional challenges, since the blue light emitted by devices can alter circadian rhythms by suppressing sleep-related brainwaves and enhancing those linked to alertness[11].

One study investigated how the COVID-19 pandemic influenced children’s screen use and sleep patterns by surveying 1,084 parents of children aged 2-18 years. Results showed that screen exposure rose significantly during the pandemic, increasing from about one hour per day to 3.5 hours (mainly for school use) and from 1.7 hours to three hours for leisure, with a marked rise in evening use after 6 p.m. Alongside this change in duration and pattern, the prevalence of sleep disturbances increased from 22.1% before the pandemic to 33.9% during and after, with the greatest increase seen in difficulties initiating and maintaining sleep[11]. Importantly, even before the pandemic, sleep problems were linked to higher screen use, especially in the evening, and the pandemic’s surge in screen time further amplified these issues. Unlike screen use that is self-paced, sporting mega-events impose externally fixed schedules, potentially amplifying circadian disruption.

Our study aims to evaluate the sleep changes during the FIFA World Cup Qatar 2022 (FWC-2022) season in children in different time zones (Saudi Arabia which shares same time zone with Qatar versus countries with a +6-hour time difference). We propose the significance of this event impact on sleep pattern among children attending FWC-2022 matches watching games at night, rather than during the evening as is the case in Saudi Arabia and other countries within the middle east time zone (+1-+2 GMT)[12]. Given that FIFA 2026 will involve an expanded format with 104 matches across three countries and different time zones[13], therefore understanding these dynamics and their effect on sleep pattern, quality and duration is increasingly urgent.

To our knowledge, while studies have examined adult sleep disruption during major tournaments, there is a paucity of data on children. This gap is critical given the global nature of FIFA events and the timing of matches across multiple time zones. By addressing this gap, our study intends to advance understanding the impact of major global events on children’s sleep patterns, which can inform guidelines for parents and educators to mitigate potential negative effects.

## 2. Methods

### 2.1. Study Design

We conducted a cross-sectional survey between 27 November and 25 December 2022, to evaluate sleep pattern changes among children during the FIFA World Cup Qatar 2022 (FWC-2022) season in different time zones (Saudi Arabia which shares same time zone with Qatar versus countries with a +6-hour time difference).

### 2.2. Sampling, Participants, and Data Collection

Survey invitations were disseminated via various social media platforms, including WhatsApp, X application (Twitter previously), and Facebook, utilizing convenience sampling methods. Participants were required to give informed consent electronically before participating in the survey. Eligible participants were parents residing in Saudi Arabia or countries with a +6-hour time difference, during the study period. The anonymity of respondents and the confidentiality of their data were guaranteed. Data collection was monitored daily and concluded once the target sample size was achieved.

### 2.3. The Survey Instrument

The survey used the validated Children’s Sleep Habits Questionnaire (CSHQ) – Arabic version [8, 9]. Demographic questions included parental gender, age groups of parents and children, and interest and methods of following the FWC-2022 matches (Appendix I). Additional questions, formulated by experts in pediatric sleep medicine and childhood health, inquired about time spent watching FWC-2022 matches, parental perception of the impact on child’s sleep, and preferences for watching football matches versus playing electronic games.

### 2.4. Sample Calculation and Recruitment

Assuming 50% of the subjects would exhibit the outcome of interest, we calculated a required sample size of 386 participants to determine the true prevalence with a 95% confidence interval and a 5% margin of error. The final sample size consisted of 848 respondents.

### 2.5. Statistical Data Analysis

Continuous variables that followed a normal distribution were described using means and standard deviations, while those that did not meet normality assumptions were described using medians and interquartile ranges. Normality was assessed using histograms and the Kolmogorov-Smirnov (KS) test. Categorical variables were described using frequencies and percentages.

Internal consistency of the sleep behaviors questionnaire was evaluated using Cronbach’s alpha. Collinearity between analyzed variables was assessed using the Variance Inflation Index (VIF) and Tolerance diagnostic tests. Multiple response dichotomies analysis was used for variables with multiple options. Bivariate Pearson’s correlations were conducted to assess correlations between metric variables.

The Children’s Sleep Behavior Questionnaire subscale scores were computed by averaging the scores of items in each subdomain according to the author’s scoring manual, with reverse scoring for specific items. A higher score indicated more problematic sleep issues.

Parental socioeconomic and educational factors were standardized into a socioeconomic factor index (SESi) using categorical factor analysis. Social jetlag (SJL) was computed by estimating the time difference between weekend and weekday sleep times, considering a delay of ≥1 hour as positive for SJL. Sleep time related jetlags due to football game days were similarly compared.

Multivariable linear regression analyses were conducted to understand the factors contributing to children’s sleep problems, assessing each sleep problem indicator (problematic bedtime routine, sleep behaviors, night waking, and morning wakeups) against sociodemographic characteristics and other sleep-related factors, accounting for social jetlag associated with weekends, weekdays, and FIFA game timings.

For dichotomous sleep-related outcomes, multivariable logistic binary regression analysis was used, with associations expressed as Odds Ratios (OR) and 95% confidence intervals. Data analysis was performed using SPSS IBM version 21, with an alpha significance level set at 0.050.

### 2.6. Ethical Considerations

The study protocol was reviewed and approved by the Institutional Review Board of King Saud University, Riyadh, Saudi Arabia (reference number: 19/0953/IRB). Participants provided voluntary electronic informed consent. They retained the right to withdraw at any stage without consequence. All data were securely stored to maintain confidentiality throughout the study.

## 3. Results

The survey results, based on a total of 848 participants, reveal that the majority were participating parents were females (60.6%) predominantly aged between 35-44 years (44.5%). Educational attainment was high, with 81.1% holding a university or postgraduate degree. 63.6% reported being employed, and the median household size was three children. A large portion of families had children aged 5-11 (70.9%) and aged 12-18 (61.2%). Regarding World Cup viewing habits, both parents (42.7%) and children (38.1%) primarily watched matches live at home. Additionally, 65.7% of respondents reported that the questionnaire prompted them to observe sleep-related symptoms in themselves or their family. Most respondents (88.6%) resided in Saudi Arabia (Table 1).

**Table-1:**
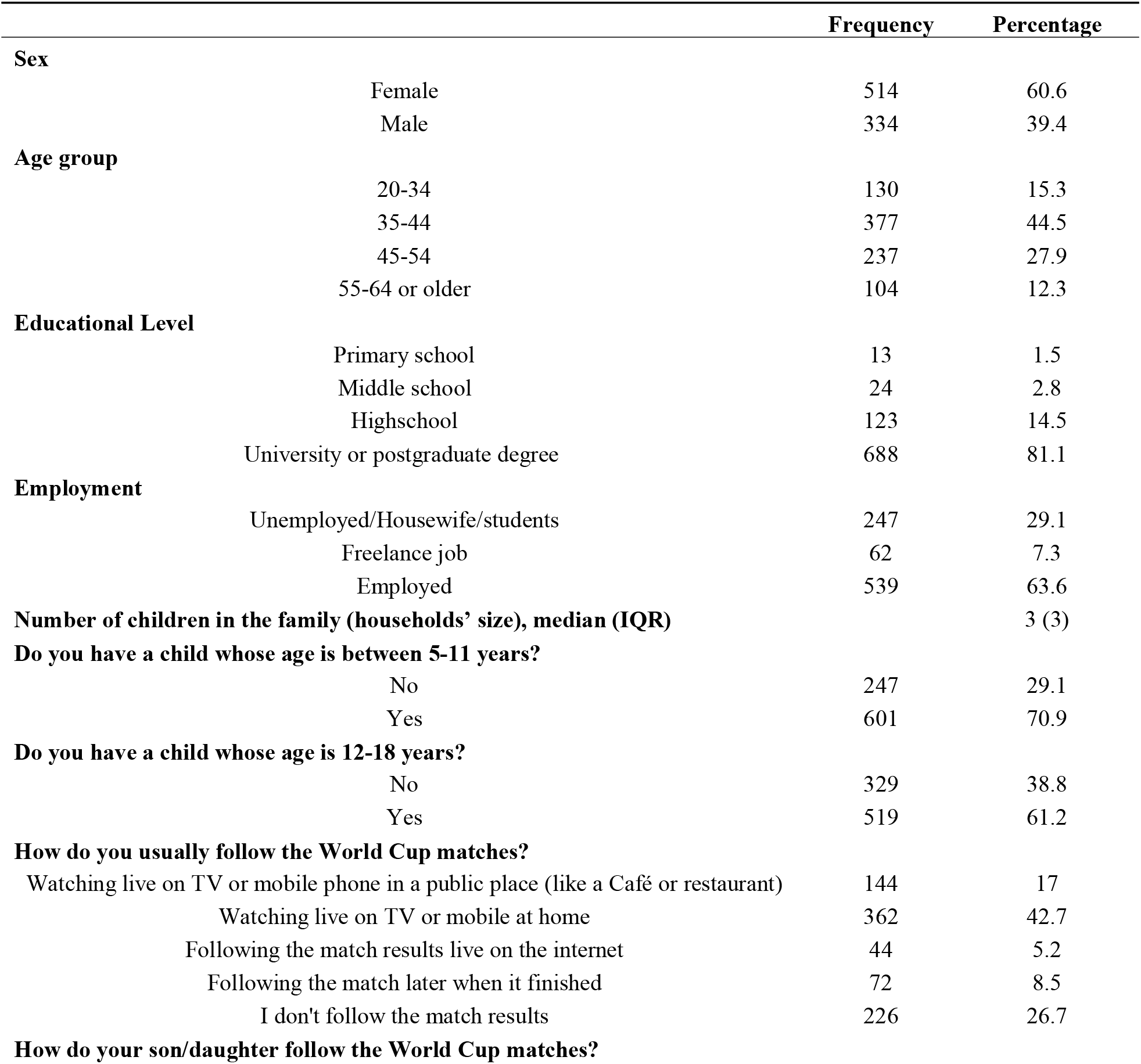
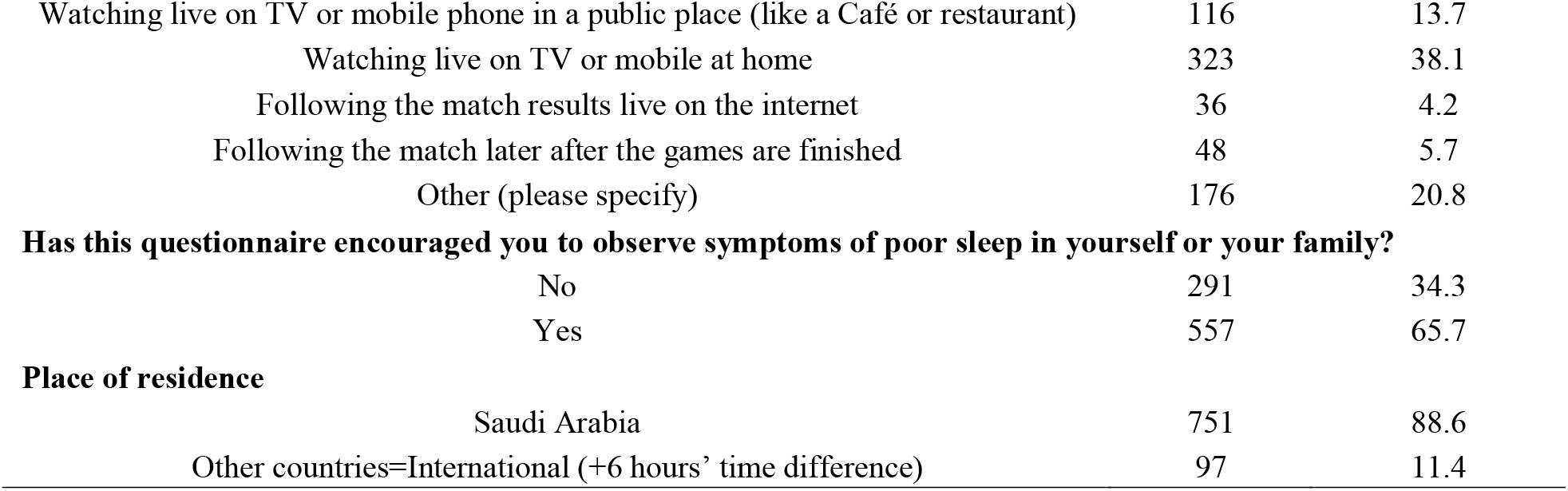
Descriptive analysis of the parents’ sociodemographic characteristics. N=848.

|  |  | Frequency | Percentage |
| --- | --- | --- | --- |
| **Sex** |  |  |  |
|  | Female | 514 | 60.6 |
|  | Male | 334 | 39.4 |
| **Age group** |  |  |  |
|  | 20-34 | 130 | 15.3 |
|  | 35-44 | 377 | 44.5 |
|  | 45-54 | 237 | 27.9 |
|  | 55-64 or older | 104 | 12.3 |
| **Educational Level** |  |  |  |
|  | Primary school | 13 | 1.5 |
|  | Middle school | 24 | 2.8 |
|  | Highschool | 123 | 14.5 |
|  | University or postgraduate degree | 688 | 81.1 |
| **Employment** |  |  |  |
|  | Unemployed/Housewife/students | 247 | 29.1 |
|  | Freelance job | 62 | 7.3 |
|  | Employed | 539 | 63.6 |
| **Number of children in the family (households' size), median (IQR)** |  |  | 3 (3) |
| **Do you have a child whose age is between 5-11 years?** |  |  |  |
|  | No | 247 | 29.1 |
|  | Yes | 601 | 70.9 |
| **Do you have a child whose age is 12-18 years?** |  |  |  |
|  | No | 329 | 38.8 |
|  | Yes | 519 | 61.2 |
| **How do you usually follow the World Cup matches?** |  |  |  |
|  | Watching live on TV or mobile phone in a public place (like a Café or restaurant) | 144 | 17 |
|  | Watching live on TV or mobile at home | 362 | 42.7 |
|  | Following the match results live on the internet | 44 | 5.2 |
|  | Following the match later when it finished | 72 | 8.5 |
|  | I don't follow the match results | 226 | 26.7 |
| **How do your son/daughter follow the World Cup matches?** |  |  |  |

|  |  |  |
| --- | --- | --- |
| Watching live on TV or mobile phone in a public place (like a Café or restaurant) | 116 | 13.7 |
| Watching live on TV or mobile at home | 323 | 38.1 |
| Following the match results live on the internet | 36 | 4.2 |
| Following the match later after the games are finished | 48 | 5.7 |
| Other (please specify) | 176 | 20.8 |
| **Has this questionnaire encouraged you to observe symptoms of poor sleep in yourself or your family?** |  |  |
| No | 291 | 34.3 |
| Yes | 557 | 65.7 |
| **Place of residence** |  |  |
| Saudi Arabia | 751 | 88.6 |
| Other countries=International (+6 hours' time difference) | 97 | 11.4 |

Table A1 describes the parents’ perceptions of their children’s sleep according to (CSHQ). Major bedtime challenges included needing rocking to fall asleep, difficulty sleeping alone, and requiring comfort objects. For sleep behaviors, the most problematic issues are night awakenings with distress, teeth grinding, loud snoring, and bed-sharing. Other major concerns were inconsistent sleep duration and daytime napping. Nighttime wakeups often involve waking more than once, while morning issues include falling asleep during activities, daytime fatigue, early waking, and difficulties with self-waking (Table A1).

The descriptive analysis of the parents’ perceptions of their children’s problematic sleep habits (Table 2) showed elevated scores across all domains of the CSHQ. The global CSHQ score was 62.19 ± 10.6, which exceeded the established clinical cutoff of 41, indicating that most children were perceived to have clinically significant sleep problems. Among the subscales, problematic sleep behaviors had the highest relative burden, with a mean score of 20.41 out of 28 points, ranking first. This was followed by bedtime routine difficulties with a mean score of 25.48 out of 36 points, ranking second. Night waking issues were also notable, with a mean score of 5.52 out of 8 points, ranking third, while morning wake-up difficulties were comparatively less prominent, with a mean score of 10.78 out of 16 points, ranking fourth. These results suggest that parents most often reported challenges with children’s sleep behaviors and bedtime routines, followed by night wakings and morning wake-up problems.

**Table-2:**
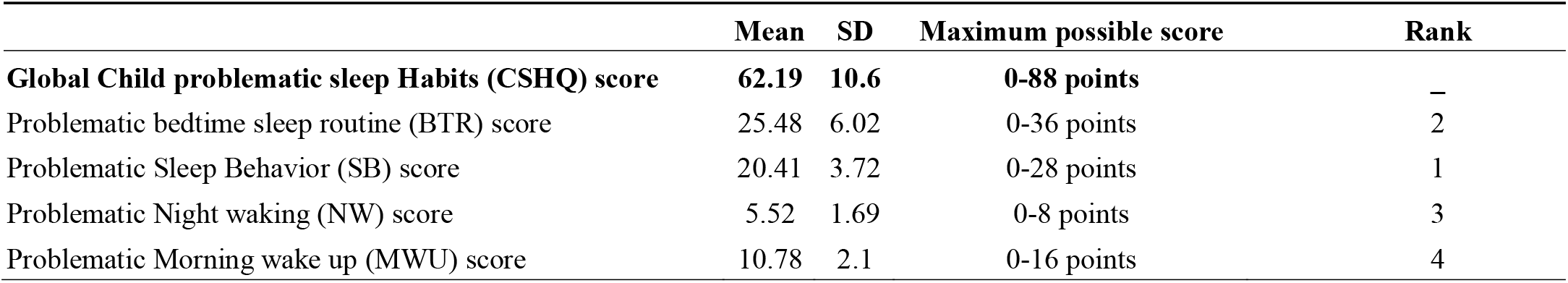
Descriptive analysis of the parents’ overall perceptions of their children’s problematic sleep habits subscale scores.

|  | Mean | SD | Maximum possible score | Rank |
| --- | --- | --- | --- | --- |
| **Global Child problematic sleep Habits (CSHQ) score** | **62.19** | **10.6** | **0-88 points** | – |
| Problematic bedtime sleep routine (BTR) score | 25.48 | 6.02 | 0-36 points | 2 |
| Problematic Sleep Behavior (SB) score | 20.41 | 3.72 | 0-28 points | 1 |
| Problematic Night waking (NW) score | 5.52 | 1.69 | 0-8 points | 3 |
| Problematic Morning wake up (MWU) score | 10.78 | 2.1 | 0-16 points | 4 |

**Figure 1:**
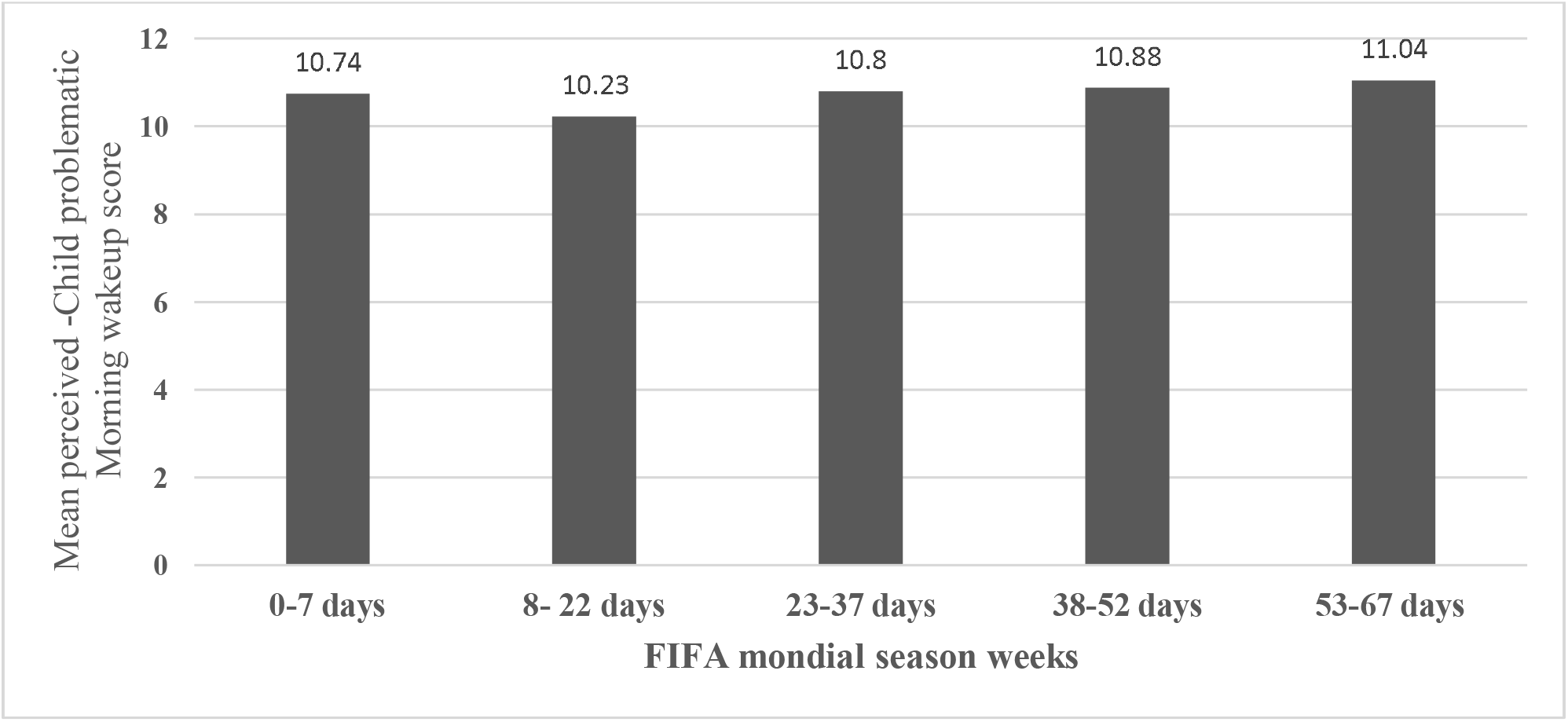
The parental Mean perceived Child problematic Morning wakeup score throughout the Mondial season.

**Figure 2:**
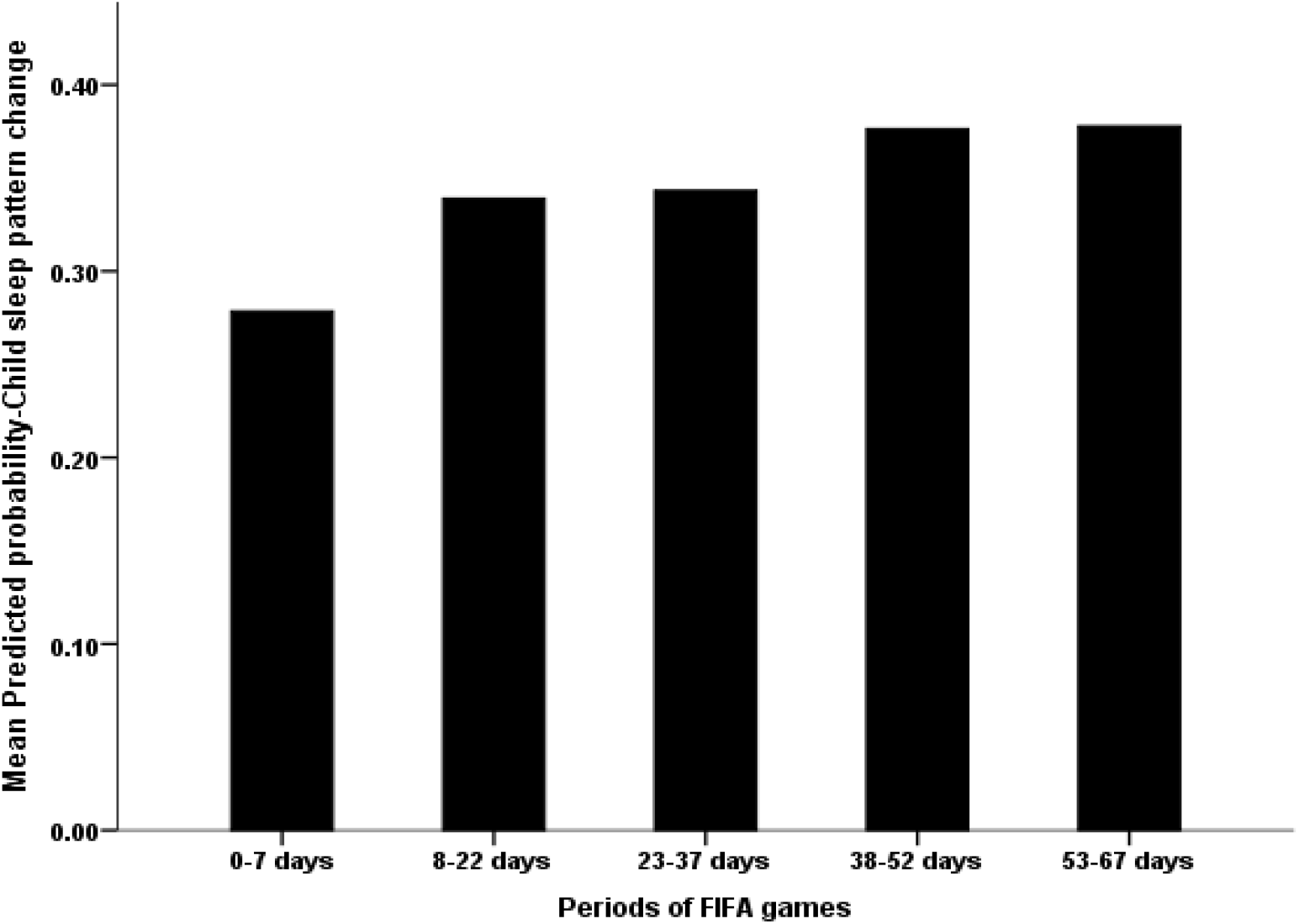
The Association Between FIFA World Cup Season Days and the Parental Predicted Probability of Child Sleep Pattern Change.

The multivariable linear regression analysis (Table 3) identified several key factors influencing parental perceptions of children’s problematic bedtime routines. Male parents perceived their children’s bedtime routines as significantly more problematic compared to female parents (β = 0.943, p = 0.017). Parents aged 45 or older reported significantly more problematic bedtime routines of their children compared to younger parents (β = 0.689, p < 0.001). Children’s problematic sleep behavior scores (β = 0.820, p < 0.001) and night waking scores (β = 0.527, p < 0.001) were significantly associated with higher problematic bedtime routine scores. The household socioeconomic status index (SESi) correlated negatively with children’s problematic bedtime routine scores (β = -0.568, p = 0.003). Parents in +6 hours’ time zone perceived significantly fewer problematic bedtime routines compared to Saudi parents in Saudi Arabia (β = -2.029, p < 0.001).

**Table-3:** Multivariable Linear Regression Analysis of the children’s bedtime sleep routine score.

| | Unstandardized<br>Beta ($\beta$)<br>Coefficients | 95% CI for Beta<br>coefficient | | p-value |
| --- | --- | --- | --- | --- |
|  |  | Lower<br>Bound | Upper<br>Bound |  |
| (Constant) | 3.411 | .833 | 5.988 | .010 |
| Sex of the parent= Male | .943 | .169 | 1.717 | .017 |
| Age $\geq 45$ years | .689 | .330 | 1.049 | <0.001 |
| Children's Sleep Behavior score | .820 | .720 | .921 | <0.001 |
| Children's night waking score | .527 | .312 | .742 | <0.001 |
| Children's morning wake-up score | .142 | -.017 | .301 | .081 |
| Households' socioeconomic status (SESI) score | -.568 | -.939 | -.198 | .003 |
| Location=International (+6 hours from Qatar and KSA region) | -2.029 | -3.019 | -1.039 | <0.001 |
| Measured delayed sleep time during weekdays due to FIFA= Yes | .087 | -.718 | .892 | .832 |
| Measured delayed sleep time during weekends due to FIFA= Yes | -.235 | -1.126 | .656 | .605 |

*Dependent outcome variable= problematic Child bedtime routine (BTR) score. Model overall significance: $f(10, 837) = 62.22$ , $p$ -value<0.001. Model $r$ -squared=0.426, adj. $r$ -squared=0.420.*

The multivariable linear regression analysis (Table 4) revealed several key findings regarding factors associated with parental perceptions of child sleep behaviors during FIFA season (SB). Children’s bedtime routines, night-time wake-ups, and morning wake-up difficulties all positively correlated with higher SB scores, with beta coefficients of 0.287, 0.292, and 0.288, respectively (all p<0.001). interestingly, international parents rated their children’s SB scores significantly higher than local parents (beta coefficient=0.659, p=0.028).

**Table-4:**
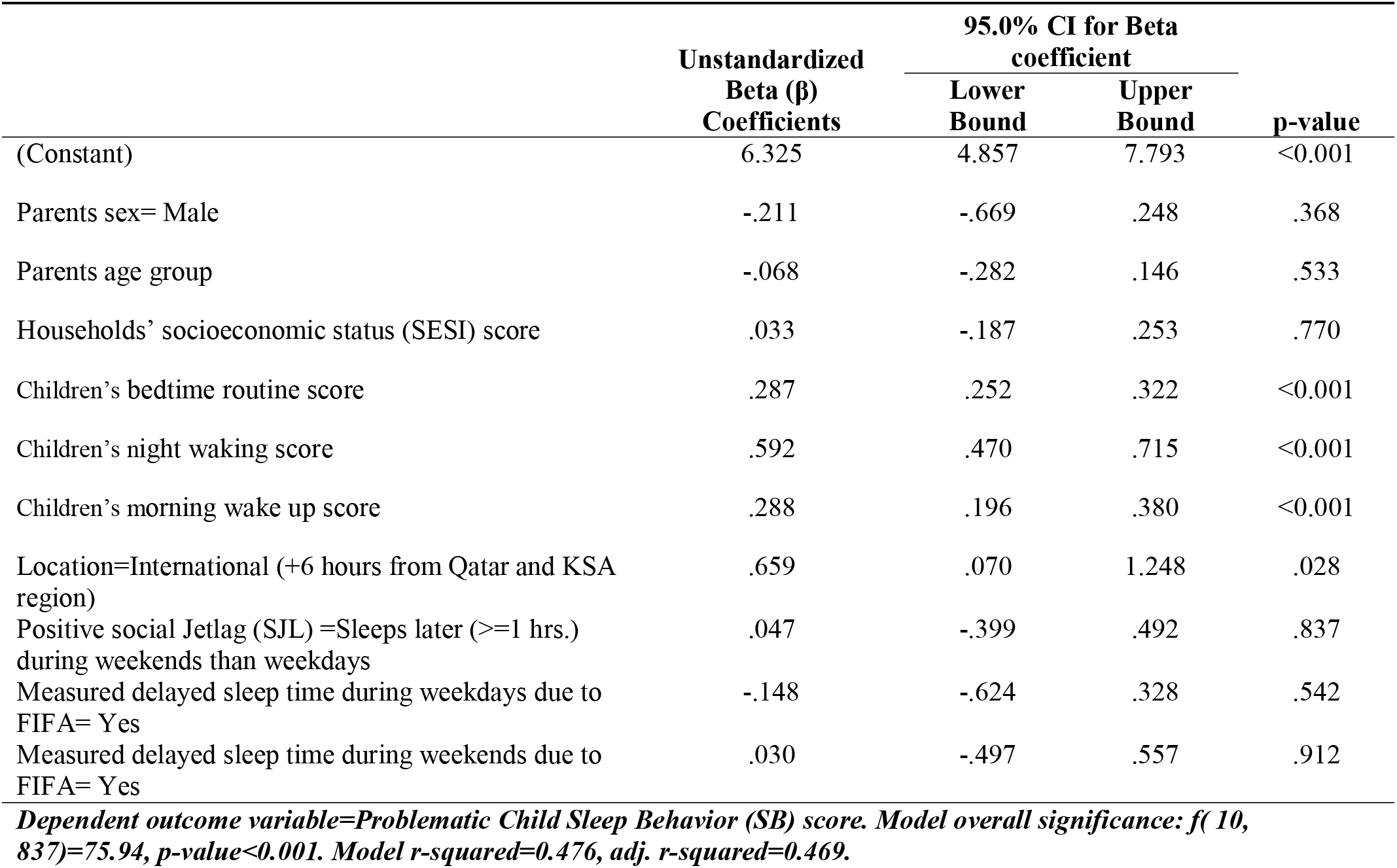
Multivariable Linear Regression Analysis of the children’s sleep behaviors score.

| | Unstandardized<br>Beta ($\beta$)<br>Coefficients | 95.0% CI for Beta<br>coefficient | | p-value |
| --- | --- | --- | --- | --- |
|  |  | Lower<br>Bound | Upper<br>Bound |  |
| (Constant) | 6.325 | 4.857 | 7.793 | <0.001 |
| Parents sex= Male | -.211 | -.669 | .248 | .368 |
| Parents age group | -.068 | -.282 | .146 | .533 |
| Households' socioeconomic status (SESI) score | .033 | -.187 | .253 | .770 |
| Children's bedtime routine score | .287 | .252 | .322 | <0.001 |
| Children's night waking score | .592 | .470 | .715 | <0.001 |
| Children's morning wake up score | .288 | .196 | .380 | <0.001 |
| Location=International (+6 hours from Qatar and KSA region) | .659 | .070 | 1.248 | .028 |
| Positive social Jetlag (SJL) =Sleeps later ($\geq 1$ hrs.) during weekends than weekdays | .047 | -.399 | .492 | .837 |
| Measured delayed sleep time during weekdays due to FIFA= Yes | -.148 | -.624 | .328 | .542 |
| Measured delayed sleep time during weekends due to FIFA= Yes | .030 | -.497 | .557 | .912 |

*Dependent outcome variable=Problematic Child Sleep Behavior (SB) score. Model overall significance: $f(10, 837)=75.94$ , $p\text{-value}<0.001$ . Model $r\text{-squared}=0.476$ , $\text{adj. } r\text{-squared}=0.469$ .*

The multivariate linear regression analysis of children’s morning wake-up (MWU) scores revealed several key findings (Table 5). Parents aged 45 years or older rated their children’s MWU scores significantly lower than parents under 45 (beta coefficient= - 0.171, p=0.032). Both children’s sleep behavior (beta coefficient=0.150, p<0.001) and children’s night-waking scores (beta coefficient=0.109, p=0.017) showed positive, significant correlations with MWU scores. Interestingly, number of family members correlated negatively with MWU scores (beta coefficient=-0.117, p=0.009), while the timing of data collection relative to the FIFA timeline showed a positive correlation, with MWU scores rising as the FIFA season progressed (beta coefficient=0.074, p=0.046) (Table 5).

**Table-5:** Multivariable Linear Regression Analysis of the children’s morning waking score.

| | Unstandardized Beta ($\beta$)<br>Coefficients | 95.0% CI for Beta<br>coefficient | | p-value |
| --- | --- | --- | --- | --- |
|  |  | Lower<br>Bound | Upper<br>Bound |  |
| (Constant) | 7.023 | 5.996 | 8.049 | <0.01 |
| Sex of the parent= Male | .115 | -.214 | .443 | .493 |
| Age group of parents>=45 years | -.171 | -.327 | -.015 | .032 |
| Household's socioeconomic status (SESI) score | -.092 | -.263 | .078 | .287 |
| Location=International (+6 hours from Qatar and KSA region) | .147 | -.276 | .569 | .496 |
| Child bedtime routine score | .026 | -.002 | .054 | .064 |
| Children's sleep Behavior score | .150 | .103 | .196 | <0.001 |
| Children's night waking score | .109 | .020 | .199 | .017 |
| Positive for social jetlag: Sleeps later (>=1 hrs.) during weekends than weekdays positive jetlag | -.055 | -.365 | .256 | .729 |
| Measured delayed sleep time during weekdays due to FIFA= Yes | -.195 | -.526 | .137 | .249 |
| Measured delayed sleep time during weekends due to FIFA= Yes | .087 | -.280 | .455 | .641 |
| Number of children in the family | -.117 | -.205 | -.029 | .009 |
| Season of FIFA games (time) =4th and later weeks | .074 | .001 | .146 | .046 |

**Dependent outcome variable=Child problematic Morning time wakeup (MWU) score. Model overall significance: $f(12, 829)=13.50$ , $p\text{-value}<0.001$ . Model $r\text{-squared}=0.163$ , $\text{adj. } r\text{-squared}=0.151$ .**

The multivariable linear regression analysis of the parental overall child sleep habits (CSHQ) score revealed several significant findings (Table 6). Parents aged 45 years or older reported significantly higher CSHQ scores compared to those younger (beta coefficient=1.473, p=0.001). Number of children within the family was associated with lower perceived sleep difficulties (beta coefficient= - 0.541, p=0.047). Social jetlag (SJL) correlated positively with higher CSHQ scores (beta coefficient=1.937, p=0.027). Parents who noticed a change in their child’s sleep pattern due to the FIFA reported lower CSHQ scores (beta coefficient= - 2.872, p<0.001). Finally, parents with children aged 12 years or older perceived significantly higher sleep difficulties compared to those with younger children (beta coefficient=2.540, p=0.004).

**Table-6:** Multivariable Linear Regression Analysis of the children’s overall sleep (CSHQ) scale score.

| | Unstandardized Beta ($\beta$)<br>Coefficients | 95.0% CI for Beta coefficient | | p-value |
| --- | --- | --- | --- | --- |
|  |  | Lower Bound | Upper Bound |  |
| (Constant) | 60.588 | 56.474 | 64.703 | <0.001 |
| Sex= Male | .923 | -.877 | 2.723 | .314 |
| Age of the parent>=45 years | 1.473 | .581 | 2.364 | .001 |
| Households' socioeconomic status (SESI) score | -.481 | -1.395 | .432 | .302 |
| Location=International (+6 hours from Qatar and KSA region) | -0.952 | -3.198 | 1.294 | 0.406 |
| Number of children in the family | -0.541 | -1.074 | -0.008 | .047 |
| Positive for social jetlag: Sleeps later ($\geq 1$ hrs.) during weekends than weekdays positive jetlag | 1.930 | 0.224 | 3.635 | .027 |
| Measured delayed sleep time during weekdays due to FIFA= Yes | -.065 | -1.893 | 1.764 | .945 |
| Measured delayed sleep time during weekends due to FIFA= Yes | -.701 | -2.716 | 1.314 | .495 |
| Perceived change level in children's sleep pattern = positively changed sleep | -2.872 | -3.941 | -1.804 | <0.001 |
| Having children aged 5-11 years | 1.430 | -.191 | 3.051 | .084 |
| Having children aged $\geq 12$ years | 2.540 | .826 | 4.255 | .004 |

*Dependent variable= Children's overall problematic sleep (CSHQ) scale score. Model overall significance: $f(12, 835)=12.24$ , $p\text{-value}<0.001$ . Model $r\text{-squared}=0.15$ , $\text{adj. } r\text{-squared}=0.137$ .*

## 4. Discussion

The present study aimed to explore the impact of the FIFA World Cup on children’s sleep patterns, assessing various sociodemographic factors, parental perceptions, and specific sleep behavior indicators. The findings revealed several significant trends and associations, providing valuable insights into the intricate dynamics between large-scale events, family demographics, and children’s sleep health.

Female parents respondent comprised 60.6% of the total participants, the tendency for females to respond more to online surveys is attributed to their natural inclination to communicate, as well as their interest in sharing opinions and experiences. This aligns with previous research indicating that women are generally more likely than men to participate in surveys, particularly those conducted by mail or online, and that they often respond more promptly to invitations. Furthermore, studies have shown that although response rates have been declining overall in recent years, the gender effect on survey participation has remained consistently evident across different survey modes and topics[14]. Additionally, many female respondents expressed concerns about maintaining their children’s sleep routines, especially during football tournaments. The majority of parents held a university degree or higher, representing 81.1% of the sample, and a substantial portion were employed, making up 63.6% of the total. These characteristics suggest that the study sample was relatively well-educated and socioeconomically stable. Previous research has demonstrated that higher educational attainment and stable employment are associated with better health outcomes, including sleep quality, for both parents and children[15].

A systematic review of experimental evidence on the relationship between sleep and video games indicates that playing video games for extended periods, especially in the evening, is likely to cause sleep problems in children. The same study also found that video games can negatively impact cognitive function on following waking days[16]. There is a correlation between delayed sleep times and increased screen time, including watching television or playing video games, particularly in the afternoon. Studies utilizing positron emission tomography (PET) scans have shown a significant release of neurotransmitters in the brain, primarily dopamine and noradrenaline, during video game play[16, 17].

Additionally, the emotional arousal associated with sports, whether for players or spectators, can make it challenging to wind down quickly and decrease the ability to fall asleep. The excitement of watching a football match combined with highly caffeinated beverages and high-sugar snacks, for example, triggers the release of adrenaline and other hormones. Adrenaline, a fight-or-flight hormone, is released in response to stressful, exciting, dangerous situations.

There is an interesting study on Spanish soccer competitive spectators who had an increase in cortisol levels during the final soccer match of the 2010 FIFA World Cup compared to those found on a control day[18]. The highest levels were observed among the most dedicated fans during the 2014 FIFA World Cup in Natal, Brazil, and there was a correlation between match outcomes and cortisol levels, with watching a loss being associated with particularly high cortisol concentrations[19].

The analysis revealed that during the 2022 FIFA World Cup period, 33% of children experienced a delay in bedtime of one hour or more on weekdays, and 17.8% on weekends. Despite these delays, 64.2% of parents observed no change in their children’s sleep hours, while 25.5% noticed a slight change. The parental perception of the World Cup’s impact on sleep varied, with 66.3% believing it had no effect, whereas only 2.2% perceived moderate to high impacts. This variability highlights the subjective nature of parental reporting and the diverse ways in which external events can influence family routines[20].

The study identified several common sleep problems among children, including needing to be rocked to sleep, difficulties sleeping alone, and requiring a transitional object. Nighttime behaviors such as sweating, screaming, and teeth grinding were also prevalent. These findings align with previous literature, which has documented similar sleep issues in children, particularly in response to environmental and lifestyle factors[21]. The Children’s Sleep Habits Questionnaire (CSHQ) scores indicated a high prevalence of problematic sleep, with a global mean score of 62.19 out of 88 points.

A study conducted in Turkey involving 114 primary school children aged 6 to 10 years revealed some significant findings regarding video game usage and sleeping habits. Among the participants, 75.9% (n=107) reported that they played video games. Alarmingly, 94.4% of those who played video games exhibited serious sleep problems, as indicated by (CSHQ) score above 41 points. Further analysis, including total scores and subgroup assessments, showed statistically significant differences in various aspects of sleep. These included total sleep score, night waking, sleep-disordered breathing, and daytime sleepiness, with p-values of less than 0.001, 0.015, 0.010, and less than 0.001, respectively[22].

Multivariable analyses revealed that male parents and those aged 45 years or older perceived their children’s bedtime routines as significantly more problematic. Higher problematic sleep behavior and night waking scores were also strongly associated with more severe bedtime issues. Additionally, higher socioeconomic status was linked to fewer bedtime routine problems, suggesting that financial stability and related resources may mitigate sleep disturbances[23]. It was also observed that international parents reported fewer problematic routines, which could be attributed to cultural differences in sleep practices and perceptions, as well as variations in time zones and demanding work and lifestyle schedules.

Bivariate correlations indicated that longer sleep duration was linked to fewer night waking problems, while having more children in the family was associated with less sleep. The regression analysis further identified key predictors of problematic sleep habits, including parental age, number of children, and changes in sleep patterns during the World Cup. Notably, parents of children aged 12 years or older reported higher problematic sleep scores, highlighting the unique sleep challenges faced by adolescents[24]. These challenges are important to address, as insufficient sleep during adolescence can be influenced by both endogenous and environmental factors. Endogenously, alterations in sleep regulation, including the circadian timing system and the sleep/wake homeostatic system, can increase physiological, cognitive, and emotional arousal, reducing sleep readiness. Environmentally, evening exposure to bright light, particularly short-wavelength blue light from electronic devices, can disrupt circadian rhythms by suppressing melatonin release, which is closely associated with sleep timing and propensity. Such exposure can delay bedtimes and sleep onset, further contributing to the sleep difficulties observed[25].

Absence of sleep disturbance among children during FWC-2022 could encourage future adaptation of World Cup virtual reality (WC-VR) in sport events, that could provide encouragement to children to enjoy the following up their favorite matches without disturbing their sleep patterns[26]. Since children in particular tend to prefer electronic games, incorporating gamification and additional physical exercise could improve the child’s engagement with WC-VR, potentially benefiting both their psychological and physical well-being. New AI-powered apps, biocomputers and wearables could enable real-time monitoring of children’s sleep during global tournaments, offering parents personalized recommendations to improve children’s sleep and balanced lifestyles[27, 28]. Furthermore, predictive analytics could help broadcasters and policymakers design more child-friendly match schedules.

## 5. Limitations and Strengths

First, the cross-sectional design of this study does not allow for the inference of causal relationships or variations between watching football matches and children’s sleep patterns, as these relationships are likely multifactorial. The habits of watching football matches may change over the season, as teams’ performances and schedules evolve. Second, the generalizability of the findings may be limited to the countries where responses were gathered. Different time zones can have varying effects on sleep, especially if participants were watching live matches, which might lead to different findings if data were collected in another time zone. Third, the use of non-probability snowball sampling limits the representativeness of the sample and the generalizability of the results to the broader population. Given reliance on social media for distribution, the sample may overrepresent digitally-active parents. Fourth, although we used a validated sleep scale, the assessments of children’s sleep symptoms were based on self-report measures, introducing potential biases that may influence the results. Finally, the findings may not be generalizable to children in other countries or to future sporting events, highlighting the need for further research.

## 6. Conclusions

This study reveals that sociodemographic factors, family dynamics, and extended events like the FIFA World Cup can influence parental perceptions of child sleep issues. Older parents and those from smaller families reported more bedtime and morning wake-up challenges, while higher socioeconomic status was linked to fewer bedtime difficulties. Although direct correlations between the World Cup and sleep disturbances were minimal, parents noticed more sleep-related symptoms as the season progressed, hinting at the subtle impacts of prolonged event schedules on family routines. These findings suggest that maintaining regular routines during large-scale events may help families manage sleep disruptions more effectively. The findings emphasize the need for interventions to maintain healthy sleep patterns during major global events. One potential solution could involve utilizing technologies like World Cup virtual reality (WC-VR) to engage children in sports without disrupting their sleep. Future research should explore these impacts across different cultural and geographical contexts to inform guidelines for parents and educators.

## Data Availability

All data produced in the present study are available upon reasonable request to the authors

## Acknowledgement

The authors thank all participants in this survey, including parents, care givers and data collectors. While preparing this manuscript, the authors used ChatGPT-5 to refine the language and grammar, then the authors reviewed and edited the content as needed and they took full responsibility for the content of the submitted manuscript. We also thanks Hodhodata.com for their support in statistical analysis.

---

**Table-A1:** Descriptive analysis and mean ranking of the parents perceptions about their childrens problematic sleep habits (CSHQ) indicators.

|  | Mean | SD | Rank |
| --- | --- | --- | --- |
| **Bedtime Routine** |  |  |  |
| 1.Child goes to bed at the same time at night (R) | 2.87 | 0.86 | 4 |
| 2.Child falls asleep within 20 minutes after going to bed.(R) | 2.72 | 0.9 | 6 |
| 3.Child falls asleep alone in own bed. (R) | 3.19 | 1.19 | 2 |
| 4.Child falls asleep in parent's or sibling's bed. | 2.84 | 1.24 | 5 |
| 5.Child falls asleep with rocking or rhythmic movements. | 3.35 | 1.01 | 1 |
| 6.Child needs special object to fall asleep (doll, special blanket, stuffed animal, etc.). | 2.94 | 1.29 | 3 |
| 7.Child needs parent in the room to fall asleep. | 2.46 | 1.4 | 8 |
| 8.Child resists going to bed at bedtime. | 2.7 | 1.15 | 7 |
| 9.Child is afraid of sleeping in the dark. | 2.41 | 1.34 | 9 |
| **Sleep Behaviors** |  |  |  |
| 10. Child sleeps about the same amount each day.(R) | 2.91 | 0.7 | 7 |
| 11.Child is restless and moves a lot during sleep. | 2.29 | 1.08 | 6 |
| 12.Child moves to someone else's bed during the night (parent, sibling, etc.). | 2.89 | 1.16 | 4 |
| 13.Child grinds teeth during sleep (your dentist may have told you this). | 3.31 | 0.99 | 2 |
| 14.Child snores loudly. | 3.06 | 0.92 | 3 |
| 15.Child awakens during the night and is sweating, screaming, and inconsolable. | 3.46 | 0.75 | 1 |
| 16.Child naps during the day. | 2.5 | 1.2 | 5 |
| **Night time wakeups** |  |  |  |
| 17.Child wakes up once during the night. | 2.49 | 0.94 | 2 |
| 18.Child wakes up more than once during the night. | 3.04 | 0.97 | 1 |
| **Morning wake up habits** |  |  |  |
| 19.Child wakes up by him/herself.(R) | 2.4 | 1.13 | 4 |
| 20.Child wakes up very early in the morning (or, earlier than necessary or desired). | 2.37 | 1.02 | 3 |
| 21.Child seems tired during the daytime. | 2.75 | 0.92 | 2 |
| 22.Child falls asleep while involved in activities. | 3.25 | 0.92 | 1 |

*Items with ( R ) Symbol are positive sleep indicators that required reverse coding prior to estimating the subscale score .*

